# Capturing noroviruses circulating in the population: sewage surveillance in Guangdong, China (2013-2018)

**DOI:** 10.1101/2020.08.20.20173088

**Authors:** J. Lu, L. Fang, J.J. Peng, L.L. Zeng, H.F. Lin, Q.L. Xiong, L.N. Yi, T. Song, J.F. He, L. M. Sun, C.W. Ke, H. Li, H.Y. Zheng

## Abstract

Noroviruses (NoVs) are the leading cause of acute gastroenteritis outbreaks. The specific geographical distribution and expanding diversity of NoVs has posed a challenge to NoV surveillance and interventions. Here, we describe the long-term dynamic correlation between NoV distribution in sewage and in the local population by using high-throughput sequencing and operational taxonomic unit (OTU) analysis. The NoV viral loads in sewage were closely associated with the number of NoV outbreaks in the population. Compared with the viral distributions in outbreaks, the dominance of the newly emerged variants, such as GII.P17-GII.17 and GII.P16-GII.2, could be detected two months ahead in sewage. In addition, the dynamics of pre-epidemic variants, which were rarely detected from clinics, could be captured through sewage surveillance, thus improving our understanding of the viral origin and evolution. Our data highlight that the high-throughput environmental screening should become a critical part of the response to infectious diseases.

## Introduction

Currently, emerging infectious diseases present one of the greatest public health challenges. Human noroviruses (NoVs) are recognized as the leading cause of acute gastroenteritis (AGE) for all ages (*1*). Viral infections are estimated to be associated with 18% of diarrheal diseases and 212,000 deaths annually (*2*). Due to their high contagiousness, most cluster infections or outbreaks of NoVs have been reported in semienclosed settings such as hospitals, childcare centers, schools, and universities, leading to public panic regarding disease transmission (*3*).

Based on the sequence of the VP1 gene, NoVs can be classified into seven genogroups (GI-GVII) and further subdivided into more than 40 genotypes (*4*). Viruses from the GI, GII, and GIV genogroups are known to infect humans. Currently, there is no drug or licensed vaccine for NoV infections. Disease control has mainly relied on clinical surveillance, which informs us of the emergence and prevalence of different genotypes and their potential risks of causing global outbreaks. Since 1995, GII.4 has been identified as the predominant genotype responsible for 65-80% of NoV infections worldwide (*5, 6*). The novel GII.4 variant has emerged every 2-5 years and presented amino acid changes in five antigenic sites (*7*) associated with viral escape from herd immunity (*8, 9*). This epochal evolution pattern is regarded as the principal mechanism adopted by GII.4 to sustain its predominance and has led to successive pandemics in the global population (*10-12*). However, in the winter seasons of 2014-2015 and 2016-2017, novel variants of some genotypes, such as GII.P17-GII.17 and GII.P16-GII.2, emerged. These NoV genotypes, which were rarely identified previously, have replaced GII.4 and caused increasing AGE outbreaks in Asia-Pacific regions. The emergence of these novel variants was also observed in European countries, with a rising prevalence in 2015-2016 (*13*).

The temporal and spatial variances in NoV genetic distribution pose a significant challenge to NoV vaccine design and for early warning of NoV outbreaks, for which more extensive and timely surveillance is warranted. In this study, we developed a high-throughput method to capture NoVs in sewage. With this method, the dynamics of NoV distribution in Guangzhou sewage were investigated from 2013 to 2018. The corresponding epidemiological data of NoV outbreaks were subtracted from the clinical surveillance network to evaluate how sewage data could improve our knowledge of NoV circulation in the population, especially for early detection of novel NoV variants.

## Results

### Norovirus genotype distributions in AGE outbreaks, 2013-2018

The Guangdong provincial surveillance network was launched in 2013 for monitoring AGE outbreaks in 21 prefectural cities (*14*). In total, 327 NoV-associated outbreaks were reported between January 2013 and December 2018. The majority of these outbreaks (284 of 327, 87%) were caused by the GII genogroup, in which GII.4, GII.17, GII.2, and GII.3 were the predominant genotypes, causing 24, 75, 143 and 42 outbreaks, respectively. As previously reported, a novel variant of GII.17 (GII.P17-GII.17) (*14*) and a recombinant variant GII.2 (GII.P16-GII.2) (*15*) emerged in Guangdong in 2015 and 2017, respectively. These novel variants caused a significant increase (>4-fold) in AGE outbreaks and became predominant in the next 2 or 3 years (Fig. 1A). More importantly, the novel variant outbreaks occurred almost simultaneously in multiple cities, and the epidemic peaked in the following 1-2 months. In this context, an optimized surveillance system that could provide timely and extensive molecular surveillance data is warranted.

**Fig. 1.**
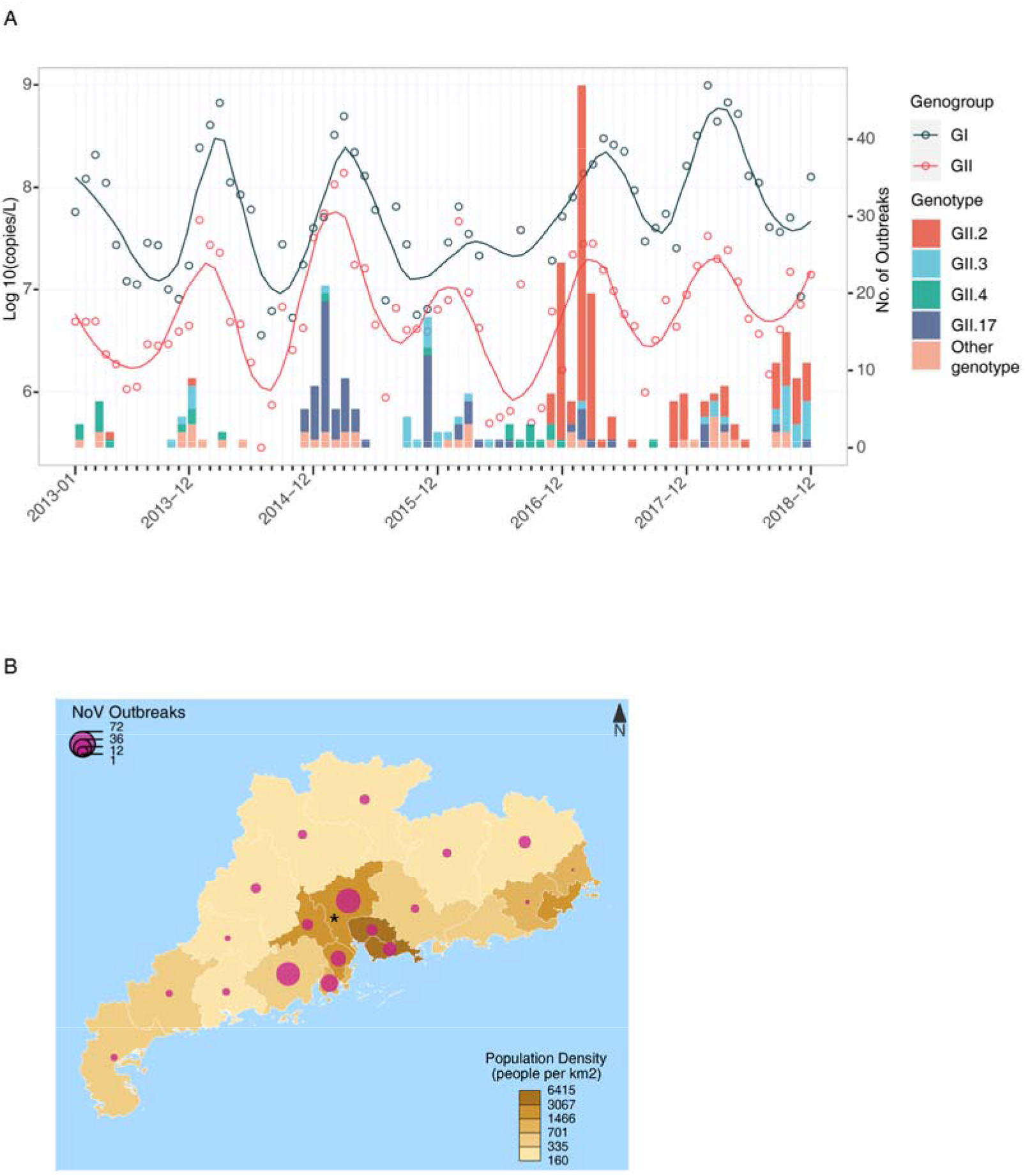
Norovirus outbreaks in Guangdong Province, China, January 2013-December 2018. (A) Temporal distribution of viral genotypes in norovirus outbreaks. The relative numbers of viral copies of GI and GII norovirus in Guangzhou sewage were plotted each month. The samples with viral loads below the detection threshold were set to a Ct-value of 40 (<1×10^5^ copies/L) for convenience. (B) Spatial distribution of norovirus outbreaks in 21 prefectural cities in Guangdong, China. Guangzhou, the hub of the norovirus epidemic, is marked with a star.

### Molecular surveillance of norovirus in raw sewage in Guangzhou, 2013-18

Guangzhou is the capital city of Guangdong Province and one of the most populous regions in China. Between 2013-2018, Guangzhou reported the largest number of NoV outbreaks (72 of 327, 22%, Fig. 1B) and was regarded as an epidemiological hub for NoV transmission (*16*). Sewage surveillance in Guangzhou was previously established for the global eradication of poliovirus and was also applied to investigate the potential origins of enterovirus outbreaks (Lu et al., 2020). To explore the dynamic correlations of NoV in sewage and in clinical outbreaks, a total of 72 sewage samples were collected from the Guangzhou wastewater treatment plant (WWTP) each month from January 2013 to December 2018. GI and GII NoVs were detected in 91.7% (66/72) and 100% (72/72) of the samples by RT-PCR. The relative virus copies in sewage samples correlated well with the number of NoV outbreaks in the population, with the most abundant GI and GII NoVs detected at the end of winter and in early spring (November to April of the next year). GI NoV was detected with relatively high viral loads in sewage but rarely identified in clinical outbreaks, suggesting that most GI infections may be asymptomatic and that these viruses circulated undetected in the local population. Although correlation was observed between the viral loads in sewage and the risks of outbreaks in the community, sewage surveillance still provided limited information for early warning of NoV outbreaks. For instance, the largest NoV outbreak was reported in the 2016-17 epidemic season, but the contemporary GII viral copies in sewage were similar to those in the other epidemic seasons. Therefore, the outbreak of NoV was not only driven by the viral prevalence in the community but could also be triggered by the emergence and transmission of a novel genetic variant for which herd immunity was lacking. Therefore, establishing a method to capture the genetic diversity of NoV in sewage is critical for optimizing surveillance.

### Setup and verification of a high-throughput method

It was expected that a sewage sample would contain different NoV genotypes and even genetic variants from a specific genotype. Capturing the distribution of multiple genotypes and variants by the conventional Sanger sequencing/cloning method was a great challenge. In this study, we used amplicon sequencing and calculated the abundances of different genotypes or variants via an operational taxonomic unit (OTU) clustering method, which is commonly used to define the taxonomic units of bacterial species in 16S rRNA sequencing. It was expected that the percentage of different genotypes or variants would be proportional to the corresponding read number in OTU clusters (Fig.2A, details in Material and Methods). To verify the accuracy of this method, we constructed four plasmids, including the genome fragments of the GII.2, GII.3, GII.4, and GII.17 genotypes. The copy numbers of these plasmids were determined through mass measurements. Serial mixtures (groups 1-3) were obtained by adding the GII.2, GII.3, GII.4, and GII.17 plasmids at ratios of 1:1:1:1, 1:3:6:9, and 1:10:100:1000 (Fig. 2B). The total amount of plasmid mixture for each group was ~0.02 ng (approximately 6×10^6^ copies). These samples were first PCR amplified, sequenced with high-throughput sequencing, and analyzed as described in the Material and Methods section. As shown in Fig. 3, the sequencing results illustrated the original genotype distribution well when different genotypes were mixed at a similar ratio. When the genotype ratio was over 1:10, PCR amplification bias likely occurred, and the genotype with a lower rate may have been overrepresented (Fig. 2B). In group 3, the ratio calculated from the number of reads in each genotype cluster was 1:6:64:246, which was basically at the same magnitude in which the plasmids were mixed (1:10:100:1000). The plasmid with the lowest copies (6×10^3^ copies) finally had 1629 clustered reads. These data suggested that the combination of high-throughput sequencing and the OTU clustering method could roughly identify the percentages of different genotypes in sewage without losing sensitivity for detecting low-proportion genotypes.

**Fig. 2.**
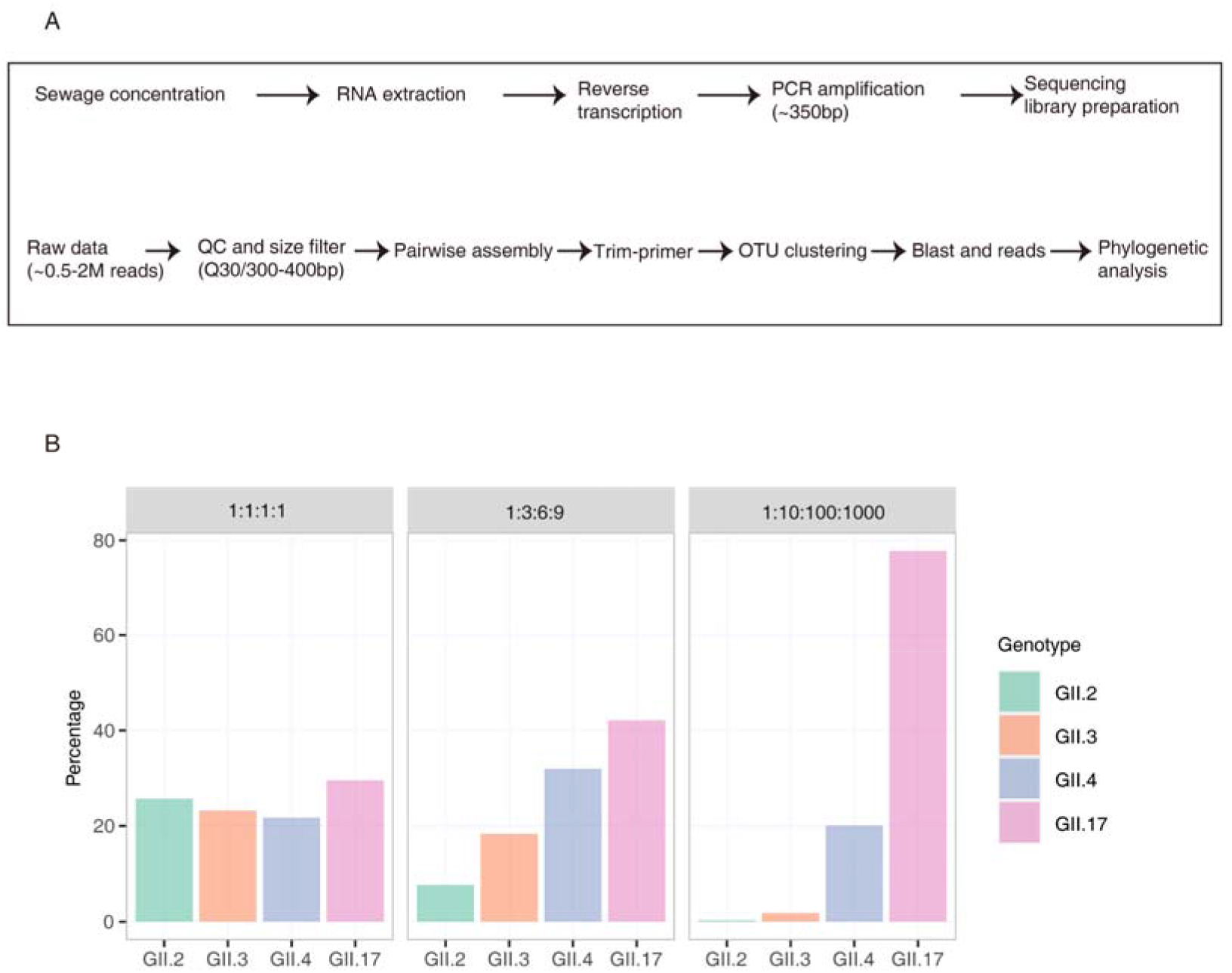
A high-throughput method for detecting NoVs in sewage. (A) workflow for calculating genotype distribution in sewage. (B) Method verification by using GII.2, GII.3, GII.4, and GII.17 plasmid constructs mixed in different ratios.

### Norovirus genotype distributions in sewage, 2013-2018

We next applied the above method to investigate NoV distribution in Guangzhou sewage from 2013-2018. Approximately 1,000,000 reads (range 112,358-2,947,207) were generated from each amplicon (Table S2). Genotype distributions in the GI and GII genogroups were determined separately by measuring the number of reads in each genotype cluster. Genotypes with a percentage less than 0.1% of total reads were removed. In total, 9 GI genotypes and 15 GII genotypes were identified in sewage water from 2013-2018 (Fig. 3A&3C). For the GI genotype, GI.2, GI.8 and GI.9 were found to be dominant in sewage samples (percentages were over 30%) but with different temporal distributions (Fig. 3A & 3B). For the GI genogroup, GI.2 was continuously circulating in the population from 2013 through 2018 (Fig. 3A), and its predominance was significant in the 2017-2018 winter season (Fig. 3B). In contrast, the GI.8 and GI.9 genotypes were dominant in 2013 but were rarely detected in 2017 and 2018 (Fig. 3A).

**Fig. 3.**
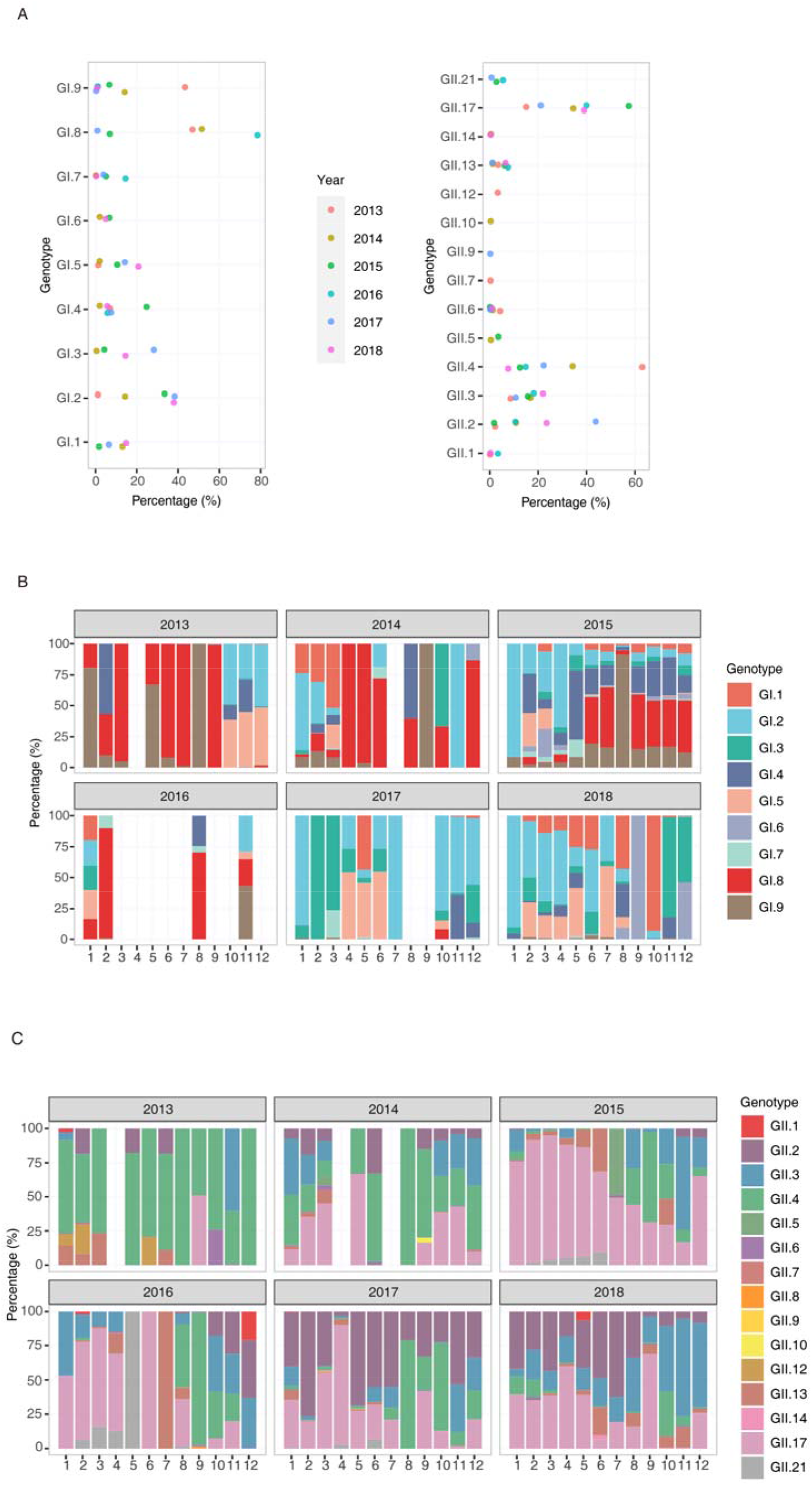
Distributions of norovirus genotypes in sewage samples. Annual distribution (A) and monthly distribution (B) of different GI genotypes in sewage samples, January 2013-December 2018. Annual distribution (C) and monthly distribution (D) of different GII genotypes, January 2013-December 2018.

GII NoV was the dominant genogroup causing AGE outbreaks. The temporal pattern of GII genotypes in sewage was well captured. GII.2, GII.4, and GII.17 were the major circulating genotypes in sewage, and other genotypes were detected with lower prevalence (<25%, Fig. 3C). More specifically, GII.4 was dominant (> 50%) in 2013 and several months of 2014 but detected at lower level in the winter season (January to April) from 2015 through 2018. GII.17 was occasionally identified as a dominant genotype in September 2013, and increasing detection of this genotype was observed in the winter season of 2014 (Fig. 3D). The percentage of the GII.17 genotype was over 50% in nearly all months from November 2014 through April 2016, consistent with its dominance in the outbreaks (Fig. 1). GII.2 was detected at a relatively low level before mid-2016 but has been increasingly identified since October 2016. Interestingly, the high prevalence of GII.4 in sewage was only observed in months before epidemic seasons (mainly July to December), and this dominance was quickly taken over by GII.17 or GII.2 in all the epidemic seasons from 2014-2018 (Fig. 3D). These data suggested that compared to GII.4, the newly emerged variants of GII.17 and GII.2 likely have higher transmission efficiency when etiological factors are favorable.

### Phylogenetic analyses

The emergence and spread of new NoV variants always lead to an increase in NoV outbreaks in the population. One priority of sewage surveillance is to identify the early emergence of novel genetic variants and characterize their dynamic distributions. In this study, we set the sequence similarity threshold at 97% in the clustering method to distinguish the different genetic variants (see Material and Methods for details). Through this method, we obtained a consensus sequence of a genetic cluster and the number of reads in it, which was proportional to the abundance in sewage. To define these genetic clusters, we constructed maximum-likelihood (ML) trees for the GII.2, GII.17 and GII.4 genotypes by combining all consensus sequences recovered from sewage and representative variant sequences from public databases (Fig.S1&S2&S3). The 350-bp amplicon sequenced in this study represented one of the most divergent parts of the viral genome and is widely used for NoV genotyping. For GII.4, nearly all variants identified after 2013 could be classified into the GII.4_Sydeny2012 lineage (Fig. 4A, top panel). The GII.4 sequences from sewage between 2013 and 2015 were near the root of the GII.4_Sydeny2012 lineage. Notably, two distinct subclusters of GII.4_Sydeny2012 emerged in August 2016 (Fig. 4A). The GII.4_Syd12_Var1 strains started to be detected in sewage in August 2016 and were more prevalent than other GII.4 variants in the following seasons (Fig. 4A, lower panel). In addition to the emergence of GII.4_Syd12_Var1, GII.4-associated outbreaks increased in winter 2016-17 compared to the low epidemic level observed in 2014 and 2015 (Fig. 4A). Phylogenetic analysis showed that the GII.4_Syd12_Var1 strains were closely related to the GII.Pe-GII.4_Sydeny2012 identified in global through the NoroNet network(*19*). The contemporary increasing epidemic observed at the end of 2016 highlighted that further characterizing the antigenicity of GII.4_Syd12_Var1 strains and their prevalence in other regions are warranted to evaluate their epidemic risk. The GII.4_Syd12_Var2 strains were closely related to the GII.P16-GII.4_Sydeny2012 strains recently reported in Europe and America (*20-22*). GII.P16-GII.4_Sydeny2012 is associated with the emergence of the novel recombinant strains GII.P16-GII.2 detected at the end of 2016 (*23, 24*).

**Fig. 4.**
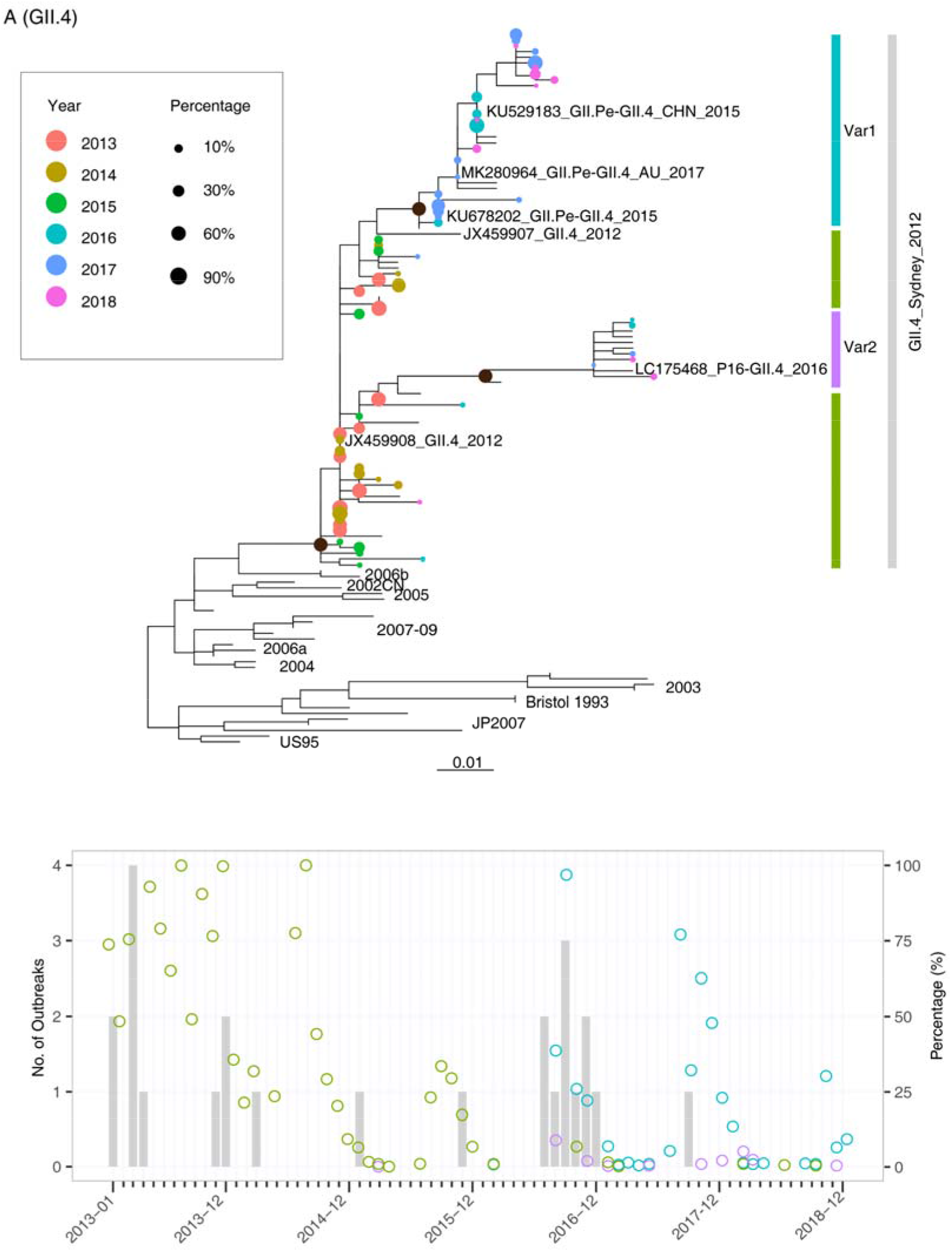
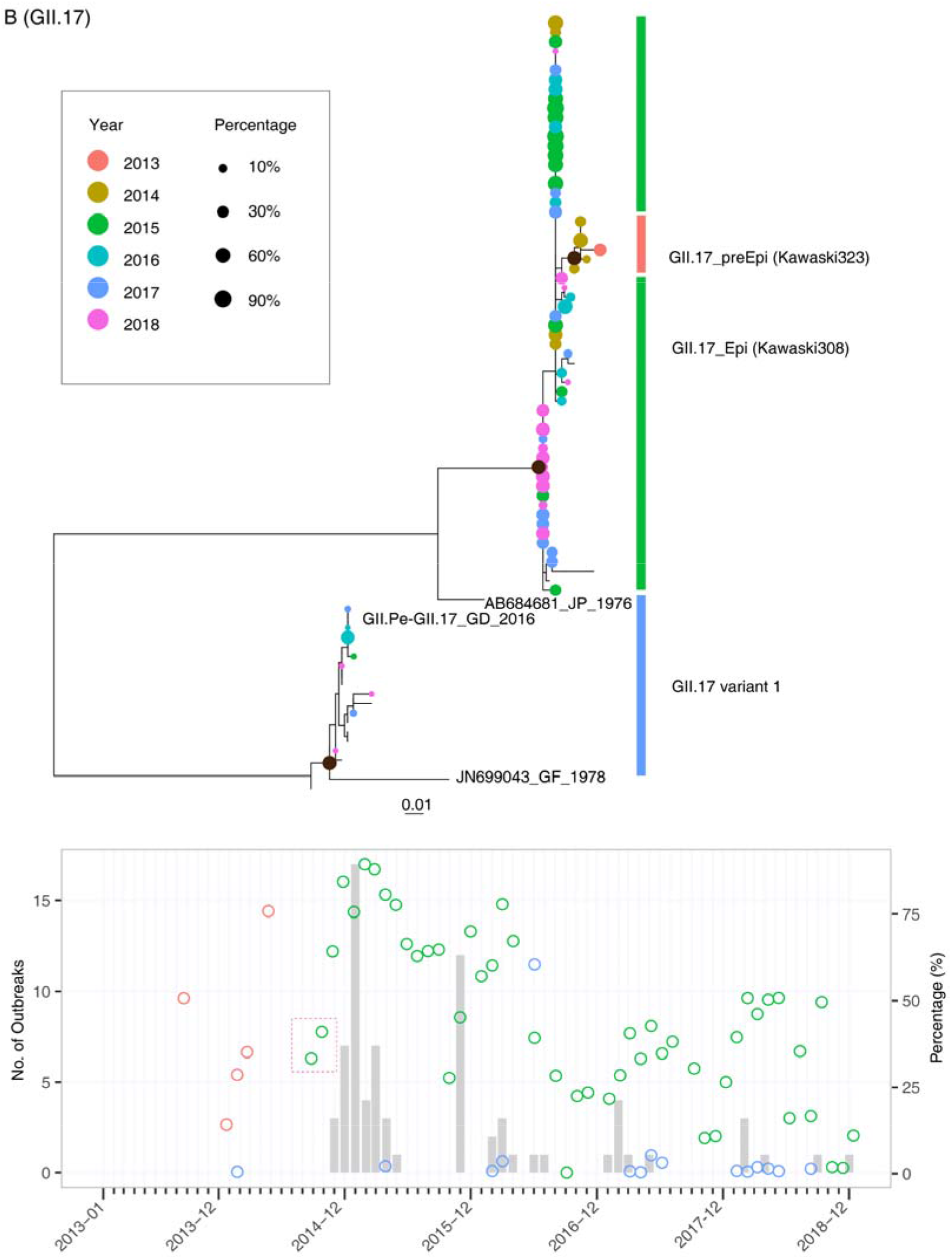
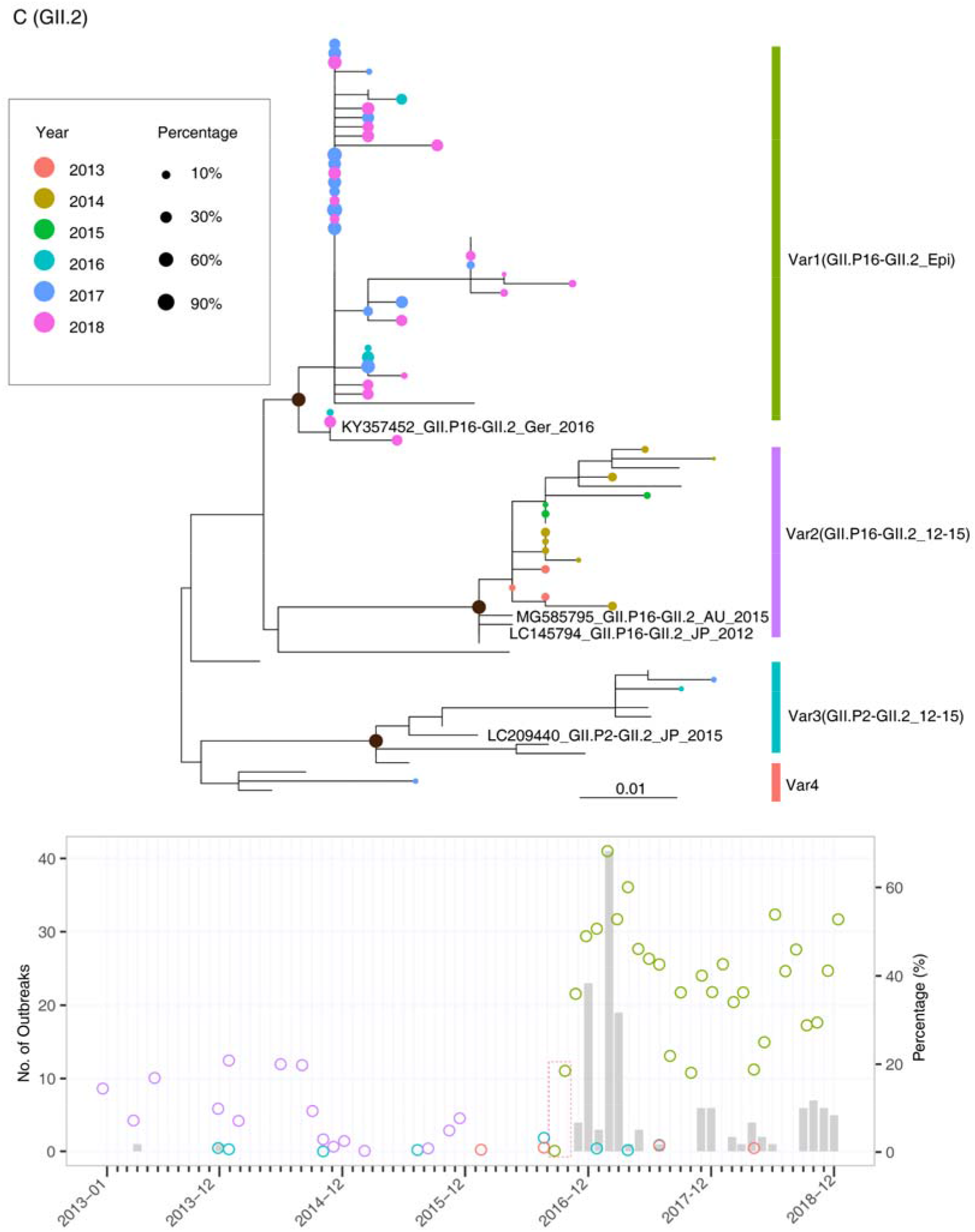
Phylogenetic analysis of different genetic variants and their dynamic changes in sewage from January 2013 to December 2018. Upper panel: maximum-likelihood trees were constructed by using the consensus sequences generated from OTU clusters of GII.4 (A), GII.17 (B), and GII.2 (C). The tip points are colored according to the year in which the samples were collected, and the size of the points was adjusted according to their abundances in sewage samples. Lower panel: The percentages of different genetic variants in sewage are plotted with circles, which are colored according to the clusters defined in the phylogenetic trees. The early detection of novel genetic variants in sewage was highlighted with a red box.

The GII.17 variant was first identified in sewage samples in September 2013, and this variant was closely related to the pre-epidemic lineage (Kawaski323-like virus) (*25*). The epidemic variant of GII.17 (Kawaski308-like virus) was first detected in sewage in September 2014, which was two months before the occurrence of AGE outbreaks. Notably, in the sewage samples, we found a GII.17 genetic variant (GII.17 variant1) that was close to the GII.Pe-GII.17 viruses recently reported in Hong Kong (KT589391_CUHK-NS-682_HK_2015-06) and southern Brazil (*26*) (Fig. 4B). This variant represented a distinct lineage originating from prototype strain JN699043_GF_1978, and it had not been identified from the outbreaks but was frequently detected in sewage between 2016 and 2018, indicative of its hidden circulation in the local community.

For GII.2, four genetic clusters were observed based on the surveillance data, and most of the GII.2 strains in sewage fell into the Var1 and Var2 clades. The Var1 clade, including the dominant strain in sewage in 2016-2018, represented the epidemic lineage GII.P16-GII.2. This lineage was regarded as a recombinant of GII.2 and GII.P16-GII.4 and caused outbreaks in multiple regions in 2016 (*23, 24, 27*). The Var2 clade included GII.2 strains mainly circulating in sewage between 2013 and 2015. Consistent with this, the closely related reference sequences for Var2 were identified in Asian-Pacific countries between 2012 and 2015 (Fig. 4C upper panel). The dynamic distribution showed that the percentage of Var2 fluctuated at a relatively low level (<25%, Fig. 4C lower panel) in sewage in 2013-2015, and this variant rarely caused outbreaks in the community. In contrast, Var1, representing the 2016 epidemic strains, was first detected in sewage in September 2016, and its percentage steadily increased in the following three months. The first occurrence of GII.P16-GII.2 in outbreak surveillance was in November 2016, which was two months after its dominance in the sewage sample.

## Discussion

Currently, multiple NoV genotypes are cocirculating worldwide with continuous and rapid changes in NoV genetic diversity (*13*). In contrast to the global predominance of GII.4 observed in the past decade, the distribution of NoV genotypes is now varied among countries, indicating that spatial and temporal differences exist. Due to this remarkable change, regional surveillance data are critical for early warning and vaccine design, similar to the actions taken in response to seasonal influenza viruses. To date, extensive and timely clinical surveillance is difficult to achieve in most regions. In this study, we prove that high-throughput sequencing combined with the OTU clustering method could be successfully applied to investigate the dynamic distribution of NoVs in sewage. By comparison with long-term surveillance data from NoV outbreaks, our results highlight that sewage surveillance serves as an efficient tool to capture the NoV distribution in the specific population.

The current international or regional NoV surveillance networks mostly focus on AGE outbreaks or sporadic infections from clinics. As a member of CaliciNet (*3*), the Guangdong provincial surveillance system followed a similar scheme to report NoV-associated AGE outbreaks in 21 prefectural cities. Great efforts have been made toward sequencing and molecular typing to capture genotype distributions, especially for the emerging novel genetic variants in the local population (*14, 15*). Several limitations have been found for the current surveillance system. The sporadic infections reported by sentinel hospitals are often biased, toward the detection of NoV genotypes associated with a relatively severe symptom. Thus, the method always oversamples specific groups of the population, such as the young or old age groups and immunocompromised individuals, who tend to have more severe infections and prolonged symptoms. Outbreak surveillance mostly identifies the genotypes that may already be efficiently transmitted in the population, and it is difficult to detect those variants before they are established in the local population. In particular, NoV genotypes such as GII.4 and GII.17 can have sudden transitions in their antigenicity or infectivity through a few amino acid mutations in their structural proteins (*28-30*). Before the transitions, the variants may have limited infectivity and circulate in the population at a relatively low level. These variants were challenging to identify during the early stage based on the conventional clinical surveillance system. For example, the pre-epidemic GII.P17_GII.17 strains had already disseminated in Guangdong in the early winter of 2013-2014 (Fig. 4B) without any associated AGE outbreaks reported. Several amino acid substitutions in the antigenic region of VP1 emerged almost simultaneously in the pre-epidemic GII.p17_GII.17 strain, resulting in the emergence of GII.p17_GII.17 epidemic strains at the end of 2014 (Fig.4B). This lineage replaced GII.4 as the predominant genotype and caused AGE outbreaks in Asian-Pacific regions in 2014-2015 (31-33). A similar pattern was also observed for GII.P16_GII.2, which was almost simultaneously identified in Germany and multiple areas in China at the end of 2016 (*15, 34*) and led to an exponential increase in NoV outbreaks (Fig. 4C). From this perspective, the time window for our response to the epidemic is too short when we rely on the current clinical surveillance system.

In this study, we discuss the possibility of capturing the NoV distribution in the local population through sewage surveillance in a hub city. Before this study, several groups tried to use Sanger sequencing/cloning or NGS to investigate NoV genotype diversity and temporal dynamics in sewage (*35-37*). However, a high-throughput method for calculating the relative abundance of different genotypes and genetic variants in sewage has not been established. The central question regarding the dynamic relationship between NoVs in sewage and the human population has still not been adequately addressed. In this study, the OTU method, which has previously been used for bacterial typing and distribution counting, was successfully applied for measuring the NoV distribution in sewage. The study results support sewage surveillance as an efficient way to capture the dynamic distribution of NoVs in the population nearly in real time (high-throughput sequencing and analyzing could be completed in 24 hours). First, the correlation between the relative viral loads in sewage and the number of outbreaks in the community suggests that the viral RNA concentration in sewage is an indicator of outbreak risks. Second, the dynamics of the novel genetic variants in sewage could be used as a warning of outbreaks in the population. For example, the epidemic strain GII.P17-GII.17 was dominant in the sewage sample in September 2014, and the increasing prevalence of this variant in October and November highlights its outbreak risk in the following epidemic season (Fig. 4B). This pattern was repeatedly observed two years later, during the outbreak of GII.P16-GII.2 (Fig. 4C). More importantly, via the clustering method, we could reveal the evolutionary history of these genetic variants, which was difficult to fully capture by clinical surveillance. As shown in Fig. 4B, the pre-epidemic GII.P17-GII.17 variant was rarely detected in clinics, possibly due to asymptomatic infections. In contrast, this variant was dominant in sewage in September 2013 and May 2014, with percentages reaching 50% and 75%, respectively. The establishment of a pre-epidemic strain in the local population provides a foundation for generating the epidemic GII.P17-GII.17 variant several months later. Similarly, the emergence and prevalence of GII.P16-GII.4-like strains identified in sewage in the middle of 2016 support the following recombination identified between GII.P16-GII.4 and GII.2 viruses.

There remain some limitations to the present study. First, specific PCR amplification is required before high-throughput sequencing, and the PCR bias would lead to low-proportion (<10%) samples being overrepresented (Fig. 2B). However, we need to balance the sensitivity and precision of this high-throughput method. The RT-PCR results showed a relatively low copy of NoVs in the raw sewage (90% samples less than 3.1×10^5^ copies/L), and direct metagenomic sequencing could obtain only a few fragments of the most abundant genotype(s). Therefore, PCR amplification is required to analyze the distribution of multiple genotypes in sewage, especially during the nonepidemic season. Second, the modified primer set G2SKF and G2SKR was used in this study to produce an ~3 50-bp fragment covering part of the VP1 region. From the sequencing data, we could not determine the RdRp genotype of NoV in sewage. Currently, it remains challenging to find a general primer set for amplifying the RdRp and VP1 fragments simultaneously, especially because this primer set should have a similar amplification efficiency among different NoV genotypes. Additionally, limited by the sequence length of the Illumina sequencing platform, the long fragment could not be sequenced without fragmentation.

For viral infectious diseases, the surveillance system is designed to answer a set of questions that are central to disease mitigation and control. These questions include what the virus is, whether it is genetically different, and how the kinetics of disease transmission are in different populations (*38, 39*). Sometimes, clinical surveillance provides a short time window for us to answer these questions and respond in an informed manner before the occurrence of large outbreaks or exponential growth of the epidemic. The global pandemic of COVID-19 is another example of how extensive clinical surveillance is needed for early warning of infectious disease epidemics. Recent sewage surveillance performed in multiple regions indicated that SARS-CoV-2 RNA could be detected in sewage before the first COVID-19 cases were reported by local clinical surveillance, and SARS-CoV-2 RNA concentration in sewage served as a leading indicator of community infection (*40, 41*). These data again highlight the importance of sewage surveillance in the early warning of emerging infectious diseases. With the significant advancement of sequencing technologies, viral disease surveillance has finally reached a point where high-throughput screening of the associated environmental samples, including sewage, air, and soil, can become a critical part of the response to infectious diseases.

## Material and Methods

### Norovirus outbreak surveillance system

The Guangdong provincial AGE outbreak surveillance network has been established since 2013 for monitoring AGE outbreaks in all 21 cities in Guangdong, China. An outbreak with a cluster of at least 20 AGE cases (meeting the Kaplan criterion) within 3 days must be reported to Guangdong Provincial Center for Disease Control and Prevention. Samples from each outbreak were first tested for NoV (Norovirus RT-PCR Kit; Shanghai ZJ Bio-Tech Co., Ltd., China) and for intestinal bacteria at the local Centers for Disease Control and Prevention. The NoV-positive specimens were delivered to the Guangdong Provincial Center for Disease Control and Prevention for further genotyping.

### Sewage surveillance

Raw sewage samples were collected monthly from January 2013 to December 2018 from the primary sedimentation tanks at the Liede wastewater treatment plant (WWTP) in Guangzhou city, China. This WWTP serves a population of approximately 2,150,000. A four-liter sample was obtained from the inlets of the primary sedimentation tanks on a routine basis each month. The sample was immediately transported to the laboratory, and sample treatment was started within 2 h after the samples arrived at the laboratory. Viruses in the sewage samples were concentrated using an improved negative-charge filter membrane absorption and sonication method, as previously described (*18*). Briefly, the collected sewage samples were centrifuged at 3,000 rpm for 30 min. Then, MgCl_2_ was added to the supernatant at a final concentration of 0.05 M, and the pH was adjusted to 3.5 to 4.0 with HCl. The samples were then slowly passed through a negatively charged membrane filter (mixed cellulose ester membrane filter; Advantec Co. Ltd., Japan) under gentle positive pressure. To elute the viruses, the filter with adsorbed viruses was cut into pieces and sonicated for 1 min in 10 ml of a 3% beef extract solution (pH 9.6), followed by centrifugation at 1,940 × g for 30 min to yield 8 ml of eluents. Finally, the eluents were passed through a 0.22 μm syringe filter to remove bacteria and fungi.

### NGS and bioinformatic analysis

The nucleic acids were extracted from 140 μl of concentrated sewage with a QIAamp Viral RNA Minikit (Qiagen, USA). RT-PCR was performed with Superscript III (Thermo Fisher, USA) by using random primers. The cDNA was then used in the first round of PCR amplification with the primer set described in Table S1. After 27 cycles of amplification, 2.5 μl of the first PCR products were used as a template in the second-round PCR. After 35 cycles of amplification, the positive products (~354 nucleotides) were subjected to Illumina MiSeq sequencing (PE250), and approximately 1,000,000 reads were generated for each PCR product. The datasets were quality controlled (QC) using fastp (*42*) to trim artificial sequences (adapters), to cut low-quality bases (quality scores < 30) and to remove short reads (< 50 bp). Paired-end reads were assembled and PCR primers were trimmed by using cutadapt. Then, the sequences were clustered into OTUs with the following clustering criteria: 1) all pairs of OTU sequences should have <97% pairwise sequence identity; 2) an OTU sequence should be the most abundant within a 97% neighborhood. These criteria have been tested to distinguish the different genetic variants of the NoV genotype, including GII.4, GII.17, and GII.2. After clustering, genotype assignment was performed using blastn (*43*) by alignment with the local NoV nucleotide database. The abundance of different OTUs and genotypes was calculated with the relative number of raw reads using R scripts. All sequencing metadata generated in this study have been submitted to the National Genomics Data Center (https://bigd.big.ac.cn/) with submission number subCRA003702.

### Phylogenetic analysis

The phylogenetic analyses were performed by combining the partial VP1 gene sequences generated from sewage samples and the selected reference sequences from the public database (Fig. S1&S2&S3) to classify the different genetic lineages. ML trees were estimated for sequences from GII.4, GII.17, and GII.2 by using IQ-Tree (*44*). Phylogenetic trees were annotated and visualized with ggtree (*45*). The dynamic changes in different genetic lineages were visualized using R scripts.

## Data Availability

All sequencing data have been submitted to public database.

## Acknowledgement

This study was supported by The Key Research and Development Program of Guangdong Province (2019B111103001) and Science and Technology Planning Project of Guangzhou (201804010030).

## Author contributions

JL designed the study. JL and HYZ had full access to all data in the study, and take responsibility for the integrity and accuracy of the data. JL, LF, JJP, LLZ, HFL and QLX contributed to NGS sequencing and the sequences analysis. JL, JJP, LLZ and QLX contributed to literature search writing of the manuscript. All authors contributed to data acquisition, data interpretation, and all reviewed and approved the final version of the manuscript.

## Supplementary Materials

**Table S1.**
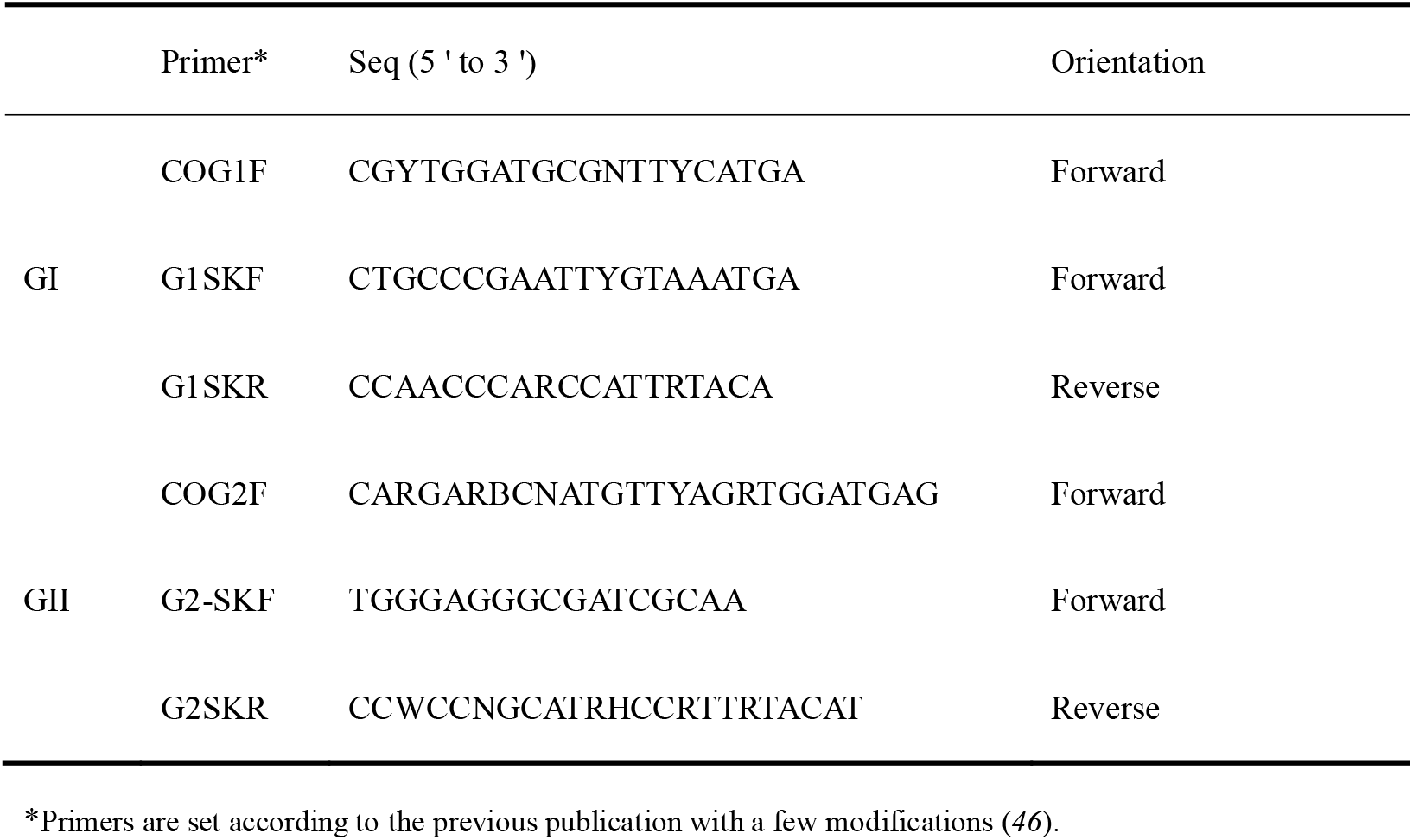
The primer set used in the PCR amplification.

**Table S2 Reads numbers of NoV collected from sewage sample in 2013-2018**

**Fig. S1. Maximum Likelihood tree of GII.4**. The accession numbers of reference sequences and the name of sequences recovered from different sewage samples have been noted. The tree was annotated with the same scheme in Fig. 4A.

**Fig. S2. Maximum Likelihood tree of GII.17**. The accession numbers of reference sequences and the name of sequences recovered from different sewage samples have been noted. The tree was annotated with the same scheme in Fig. 4B.

**Fig. S3. Maximum Likelihood tree of GII.2** The accession numbers of reference sequences and the name of sequences recovered from different sewage samples have been noted. The tree was annotated with the same scheme in Fig. 4C.

